# The Knowledge and Attitude of Physicians Regarding Vaccinations in Yerevan, Armenia: Challenges for COVID-19

**DOI:** 10.1101/2021.06.15.21258948

**Authors:** Arman R. Badalyan, Marine Hovhannisyan, Gayane Ghavalyan, Mary M. Ter-Stepanyan, Rory Cave, Jennifer Cole, Andrew W.K. Farlow, Hermine V. Mkrtchyan

## Abstract

**Background:** This primary-data analysis investigates the current level of awareness and medical knowledge of physicians in 20 health facilities in Yerevan, Armenia regarding vaccination – specifically with regard to HPV infection and the recently-introduced Gardasil vaccine used against HPV infection – that may have implications for successful roll-outs of national programmes for new vaccines, including those for COVID-19.

**Methods:** A questionnaire-based cross-sectional study was completed by 348 physicians who met the study inclusion criteria, from 20 out of 36 randomly selected healthcare facilities in Yerevan, Armenia, between Dec 2017 to Sep 2018. The aim of the questionnaire was to identify physicians’ awareness of and attitudes to HPV-related cervical cancer and the Gardasil vaccine. Responses were analysed using SPSS software (Version 16).

**Results:** The responding physicians displayed a respectable level of knowledge and awareness regarding vaccination with regard to some characteristics (e.g. more than 81% knew that HPV infection was commonly asymptomatic, 87% were knew that HPV infection was implicated in most cervical cancers and 87% knew that cervical cancer is the most prevalent cancer amongst women) but low knowledge in others and poor understanding of key issues such as the age at which women were most likely to develop cervical cancer (only 15% answered correctly); whether or not the vaccine should be administered to people who had already been affected (27% answered correctly) and whether sexually active young people should be treated for infection before vaccination (26%). Lack of confidence within the surveyed groups regarding the value of vaccination and, in particular concerns over the reasons for administering it to certain age cohorts, was driven by misconceptions.

**Conclusions:** Armenian physicians’ awareness of vaccine characteristics, the reasons for their inclusion in the national vaccination programme and the characteristics of the diseases they treat can be poor. The study further suggests that drivers of vaccine hesitancy are complex, may not be consistent from vaccine to vaccine, and may vary from generation to generation. The Armenian healthcare sector may need to provide additional training, awareness-raising and educational activities to improve understanding of and trust in vaccination programmes. Further studies are warranted to better understand knowledge, attitudes and practices (KAP) regarding immunization and vaccination programmes amongst Armenian healthcare workers.

## Introduction

Armenia has consistently displayed lower levels of vaccine confidence than surrounding countries and with Europe as a whole^1,2,3^ with Armenians showing low levels of trust in vaccines as safe, effective and/or important.^1^ This has significant implications for control and prevention of not only common endemic diseases but of emerging diseases, the frequency of which is predicted to increase during the 21^st^ century^4^, risking further pandemics such as COVID-19^5^. In this paper, we use the national vaccination programme for human papillomavirus (HPV) in Yerevan, Armenia as a case study for understanding vaccine hesitancy in Armenia more broadly, for shedding light on some key underlying reasons for hesitancy – particularly of new vaccines – and for suggesting inventions that may improve vaccine acceptance as well as confidence and thus enhance the uptake of vaccines for this and other diseases.

Human papillomaviruses (HPVs) are a group of viruses belonging to the family of *Papillomaviridae* that affect epithelial tissue, the layer or layers of cells that form the covering of most internal and external surfaces of the body and its organs. More than 150 types of HPV have been identified^6^. In women, HPV infection may cause cervical, vaginal, vulvar, anal, and oropharyngeal cancers; in males, it may cause anal, penile, and oropharyngeal cancers.^7, 8, 9^

Invasive cervical cancer (ICC) is one of the leading causes of cancer in women and, according to 2018 global estimates, results in approximately 570,000 new cases and 311,000 deaths annually^10^. In the United States, around 4,000 women die from cervical cancer each year, with African Americans and women from poorer backgrounds having a much higher rate of mortality^11^. Globally, cervical cancer is the fourth most common cancer among women. The burden faced by low- and middle-income countries is significantly higher than in high-income regions of the world^12^. The estimated age-standardised incidence of cervical cancer in 2018 averaged 13.1 per 100 000 women globally but varied from less than 2 up to 75 per 100,000 women^10^ depending on the country.

In Armenia, the cumulative incidence rate of ICC has increased from 9.5 (1985) to 16.6 (2017) per 100,000 women. Annually, about 120-130 women die from cervical cancer in Armenia (7.4/100,000 in 2016). About 50% of women with ICC are not diagnosed until the third and fourth stages of the cancer when the treatment outcome is significantly less certain^13, 14^.

Currently, there are three HPV vaccines available: bivalent (HPV 16,18), quadrivalent (HPV 6,11,16,18), and 9-valent (6,11,16,18,31,33,45,52,58) which can protect against up to nine of the most prevalent HPV genotypes associated with cancer and genital warts^15^. Of these, quadrivalent (HPV 6,11,16,18) is the most common in Armenia. Globally, HPV vaccines have been extremely successful from both a scientific and clinical viewpoint. There is strong evidence of population-level impact on the circulation of HPV infection, incidence of high-grade cervical cancer and cases of genital warts following the introduction of national vaccination programmes^16, 17^.

Despite the recommendations for routine HPV vaccinations from the World Health Organization (WHO), some countries nevertheless still remain hesitant about vaccinating their young people against HPV^18^. Armenia is one of these and the vaccine hesitancy shown with regard to HPV vaccination provides warnings of challenges that may also hinder other vaccination programmes, including COVID-19. As widespread vaccination is the most effective way to bring down cases of COVID-19 and to contain the current (and any potential future) pandemic, understanding the reasons for this vaccine hesitancy is essential if Armenia (and other countries facing the same phenomenon of vaccine hesitancy), is to avoid becoming a weak link in regional and global vaccination initiatives that are needed to bring the pandemic to an end, as well as to better control endemic diseases.^19^

Understanding the challenges of introducing a new vaccine to a hesitant population, and the underlying drivers of and for that hesitancy, is paramount. Armenia is displaying considerable vaccine hesitancy in response to the development of the COVID-19 vaccines: as of 24^th^ May 2021 only 26,562 vaccines had been administered in Armenia at a rate of around 0.8% of the population, despite ready availability of the vaccine. Conducting a study with specific reference to the COVID-19 vaccine would be ideal but, while this study was undertaken before COVID19 vaccines had been developed, using HPV vaccine as a proxy is an imperfect but practical alternative to explore how one might conceptualise vaccine hesitancy in Armenia. HPV thus offers a case study that may inform the roll-out of other national vaccination programmes.

The quadrivalent vaccine Gardasil was introduced in Armenia in December 2017^20^ but uptake has been slow, with resistance from the medical community as well as from the public. To understand possible causes of resistance from medical practitioners, we conducted a study assessing the knowledge, attitudes and practices (KAP) of physicians regarding HPV and vaccination against HPV, particularly with Gardasil, in Armenia from Dec 2017 to Sep 2018; the results provide baseline understandings of some causes of vaccine hesitancy and highlight some issues that may present similar barriers to the uptake of vaccines developed to combat COVID-19. The survey results reveal, in addition, that the term ‘vaccine hesitancy’ needs to be unpacked further as there are degrees or shades of hesitancy that range from near-acceptance to out-right rejection.

## Methods

Between December 2017 and September 2018, a questionnaire-based, cross-sectional, quantitative study was conducted in Armenian healthcare facilities to identify respondents’ awareness of and attitudes to HPV-related cervical cancer and the Gardasil vaccine. The questionnaire was adapted from a survey previously used to determine the knowledge and attitude of registered nurses towards HPV and HPV vaccine in the USA^21^. This was considered to be suitably generic that it did not require amendment to the local context. The questionnaire was administered to physicians at 20 out of 36 government-run healthcare facilities in Yerevan; the facilities included were selected at random from all those available.

The inclusion criteria for the study respondents were fully qualified medical staff from four medical specialities considered most likely to have direct interaction with HPV-infected patients or with patients eligible for HPV vaccination: paediatricians, family-practice doctors, gynaecologists and oncologists. Exclusion criteria was student doctors, nurses, residents and doctors from other specialities. This yielded a possible survey size of 405 physicians in the 20 facilities. Trained researchers from Yerevan State University School of Public Health visited each of the healthcare facilities included and personally invited physicians to participate in the study. Of the 405 eligible staff at the 20 facilities, 385 (95%) were approached to participate (the remaining 20 were not available on any of the days the researchers visited) of whom 348 (90.3%) gave consent and completed the survey. The survey was conducted by face-to-face interview in each physician’s usual place of work. The required sample size was calculated using the Estimate-proportion n=z^2^*pq/d^2^ formula.^22^ Of a total population of 405 eligible participants, 348 (90.3%) returned surveys with consent given and all sections completed. This is well above the number required to be considered a representative sample (Figure 1).

**Figure 1.**
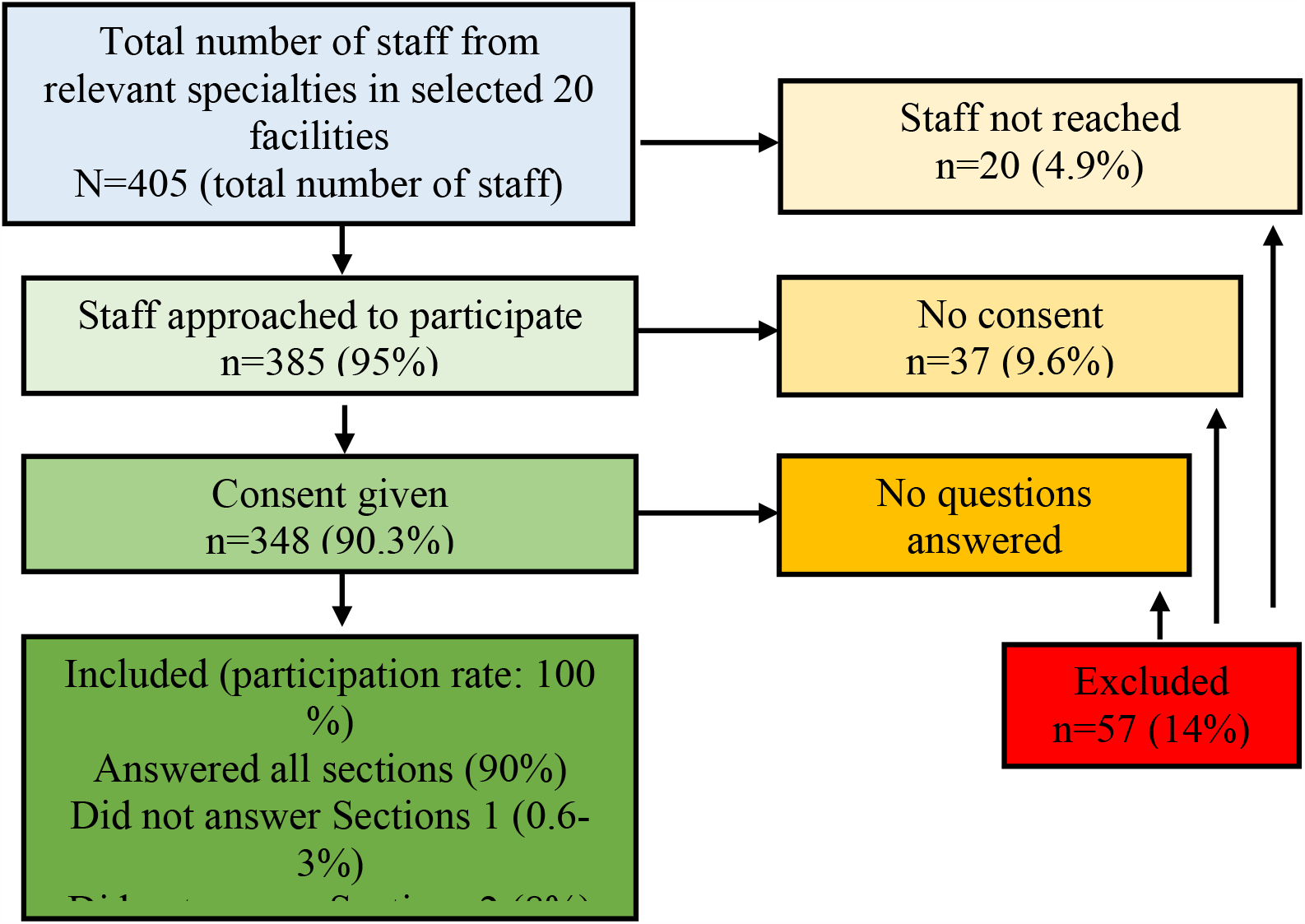
Flow chart of staff recruitment.

Data including the socio-demographic characteristics of the respondents (gender, age and length of service within the healthcare profession) were collected. The questionnaire also addressed respondents’ existing knowledge and attitudes across four areas: 1) Knowledge/Information about HPV; 2) Knowledge/Information about the HPV vaccine; 3) Attitude towards the HPV vaccine; and 4) Attitude towards other vaccines. Respondents could answer the survey questions ‘True’, ‘False’ or ‘I don’t know’, numerically coded 1, 2, and 88, respectively.

The study was approved by the Ethics Committee of Yerevan State Medical University after Mkhitar Heratsi (YSMU 14.06.2016/No 10).

### Statistical analysis

Chi-square analyses were used for the categorical variables which compared the age groups of respondents, their lengths of experience as doctors, and the fields they specialised in. For all analyses conducted in the study, an association of p<0.05 was considered to be statistically significant. Analyses were performed using SPSS software (Version 16.0).

A hierarchy heatmap on physicians’ opinions of the HPV vaccine was constructed using the R statistical computer programme language package ‘pheatmap’ (https://www.rdocumentation.org/packages/pheatmap/versions/1.0.12/topics/pheatmap) based on the percentage of respondents who answered ‘yes’ to a series of questions.

### Multivariate logistic regression analysis

Multivariate logistic regression analysis was performed to determine which factors effect awareness of HPV and its vaccine amongst physicians in Armenia. Using the glm function in R, nine logistic regression models were constructed to determine the odds ratio (OD) and 95% confidence intervals (CI) for physicians answering at random: 1) HPV is relatively uncommon 2) Almost all cervical cancers are caused by HPV 3) HPV is most common in women in their 30s 4) Cervical cancer is one of the most prevalent cancers among women 5) Most people with genital HPV are symptomatic 6) Genital warts are caused by the same HPV types that cause cervical cancer 7) Sexually active adolescents should be tested before HPV vaccination 8) HPV vaccine is available for both males and females and 9) Men and women who have been diagnosed with HPV should not be given HPV vaccine. Factors that were considered in the model were physicians’ speciality, age and experience.

### Limitations

Our study is represented by health facilities (and physicians) from Yerevan only, therefore the findings may not be generalizable to the whole of Armenia. Knowledge and attitude of respondents working in medical facilities located out of Yerevan may vary from study results. The assumptions we made about the generalisability of these findings to vaccine confidence in general and confidence to COVID-19 vaccines in particular are assumptions only and need to be tested in further research studies.

## Results

A total of 348 (90.3%) participants from the 20 randomly selected health facilities (from total of 36 health facilities located in Yerevan) completed the survey and were included in the final analysis (Figure 1).

The respondents were predominantly female (95%), reflecting the demographics of the professions (paediatricians, family-practice physicians, gynaecologists, and oncologists). The gender inequality among the respondents (95% female) is not a limitation of this study: according to recent statistics, in Armenia female doctors are more likely to specialise in paediatrics, family-practice and gynaecology, hence the high levels of female respondents in this study. A report published by the Ministry of Health, Armenia in 2016 shows that 40-60% of physicians were female and of these 91.8% were paediatricians, family-practice physicians and general practitioners, of which 95.2% were paediatricians^23^. As previous studies of vaccine hesitancy either recorded no significant differences between men and women^24^ or have been inconsistent over which gender is more likely to be vaccine hesitant^25,26^, a heavily female survey cohort is unlikely to influence perceptions of vaccine hesitancy overall. Of those participating in this study, 62% were paediatricians, 16% family-practice physicians, 13% gynaecologists, and 9% were oncologists. The age distribution was 29-60+years of age, with 52% between 45 and 59 years old. Two-thirds (67%) had more than 20 years of experience as doctors.

### Knowledge of HPV transmission, symptoms and disease

In response to some questions, study participants displayed strong awareness of the issues surrounding HPV infection and its related health outcomes, for instance most knew that cervical cancer is one of the most prevalent types of cancer among women (87.2%) and nearly three-quarters (73.4%) knew that most cervical cancers are caused by the HPV virus (73.4%). (Table 1). Awareness of presentation of the disease was also high, with 81% aware that most people with genital HPV infections are unlikely to display symptoms of the disease.

**Table 1.**
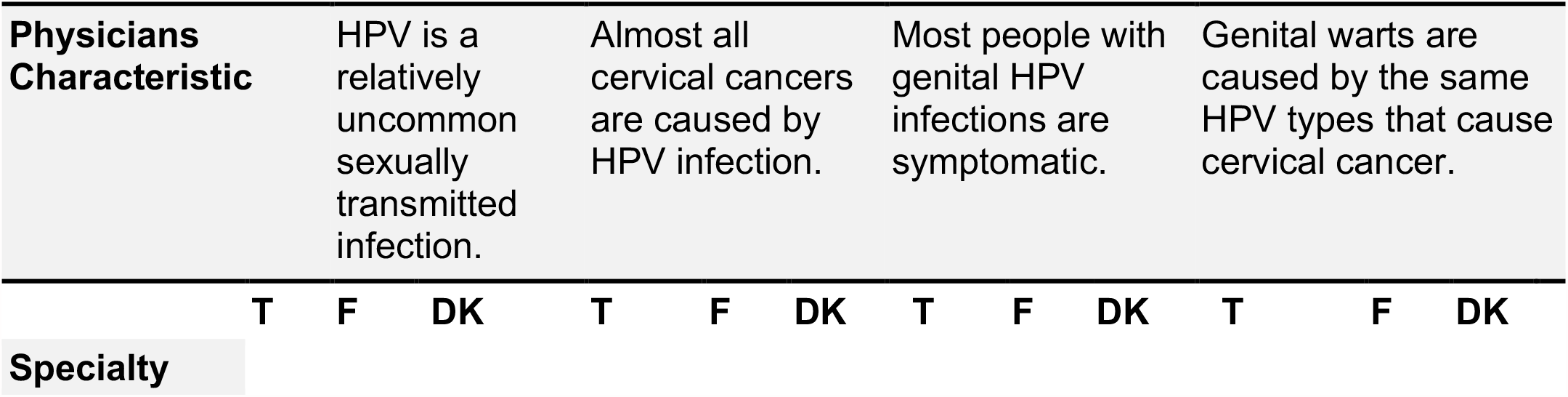
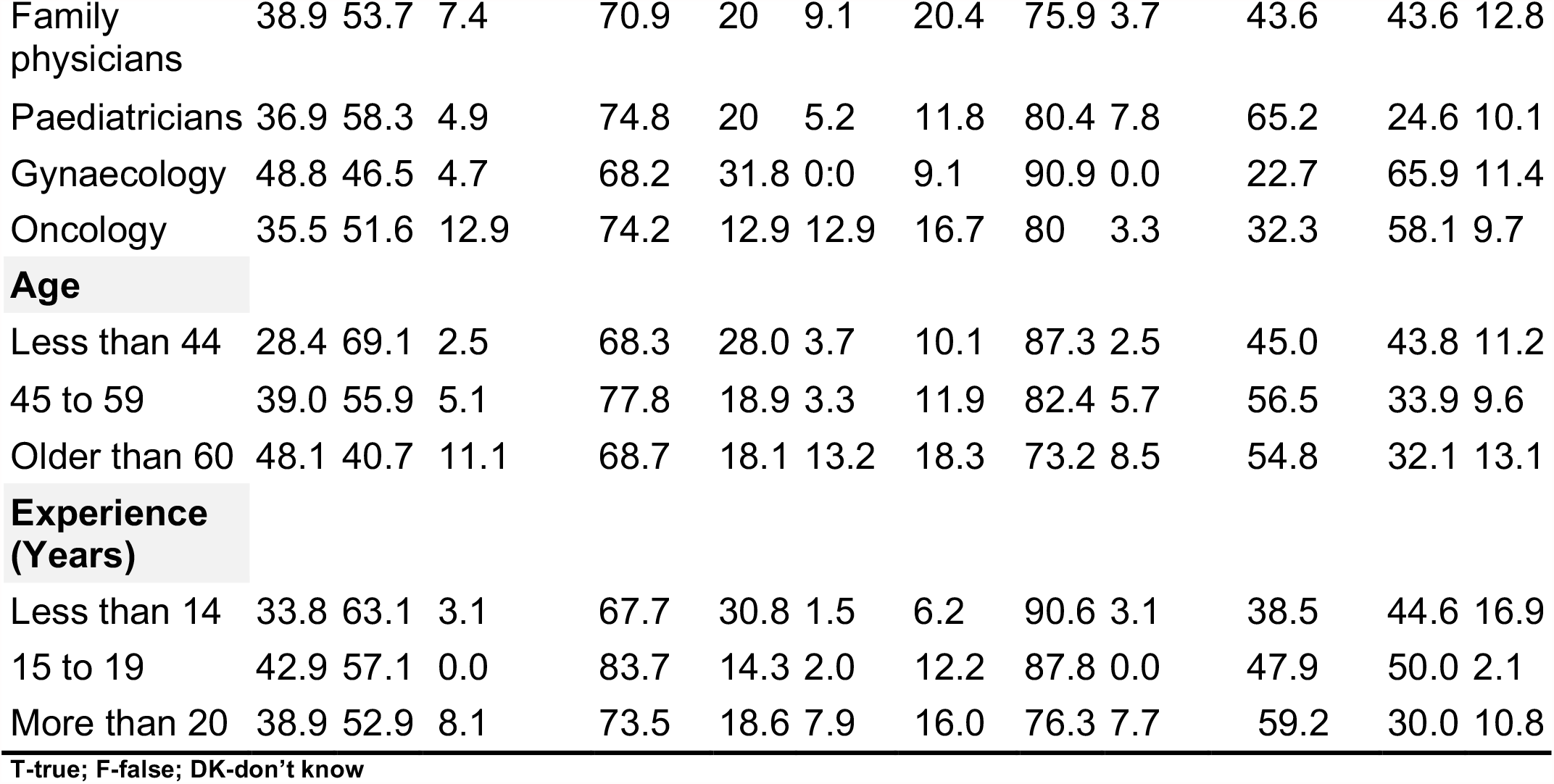
Knowledge of HPV of physicians practicing different specialist areas.

|  |  |  |  |  |  |  |  |  |  |  |  |  |
| --- | --- | --- | --- | --- | --- | --- | --- | --- | --- | --- | --- | --- |
| Family physicians | 38.9 | 53.7 | 7.4 | 70.9 | 20 | 9.1 | 20.4 | 75.9 | 3.7 | 43.6 | 43.6 | 12.8 |
| Paediatricians | 36.9 | 58.3 | 4.9 | 74.8 | 20 | 5.2 | 11.8 | 80.4 | 7.8 | 65.2 | 24.6 | 10.1 |
| Gynaecology | 48.8 | 46.5 | 4.7 | 68.2 | 31.8 | 0:0 | 9.1 | 90.9 | 0.0 | 22.7 | 65.9 | 11.4 |
| Oncology | 35.5 | 51.6 | 12.9 | 74.2 | 12.9 | 12.9 | 16.7 | 80 | 3.3 | 32.3 | 58.1 | 9.7 |
| **Age** |  |  |  |  |  |  |  |  |  |  |  |  |
| Less than 44 | 28.4 | 69.1 | 2.5 | 68.3 | 28.0 | 3.7 | 10.1 | 87.3 | 2.5 | 45.0 | 43.8 | 11.2 |
| 45 to 59 | 39.0 | 55.9 | 5.1 | 77.8 | 18.9 | 3.3 | 11.9 | 82.4 | 5.7 | 56.5 | 33.9 | 9.6 |
| Older than 60 | 48.1 | 40.7 | 11.1 | 68.7 | 18.1 | 13.2 | 18.3 | 73.2 | 8.5 | 54.8 | 32.1 | 13.1 |
| **Experience (Years)** |  |  |  |  |  |  |  |  |  |  |  |  |
| Less than 14 | 33.8 | 63.1 | 3.1 | 67.7 | 30.8 | 1.5 | 6.2 | 90.6 | 3.1 | 38.5 | 44.6 | 16.9 |
| 15 to 19 | 42.9 | 57.1 | 0.0 | 83.7 | 14.3 | 2.0 | 12.2 | 87.8 | 0.0 | 47.9 | 50.0 | 2.1 |
| More than 20 | 38.9 | 52.9 | 8.1 | 73.5 | 18.6 | 7.9 | 16.0 | 76.3 | 7.7 | 59.2 | 30.0 | 10.8 |

T-true; F-false; DK-don't know

However, 44.4% of the participants were unaware that HPV is a relatively common sexually transmitted infection and 29% of respondents unaware that the vaccine is available to both females and males. More than half (53.5%) answered that the same types of HPV that cause genital warts also cause cervical cancer, which is incorrect. Only 27% of the respondents were aware that both male and female patients diagnosed with HPV should still be encouraged to receive the vaccine, and only a similar proportion (26%) knew that sexually-active adolescents do not need to be tested before receiving their HPV vaccinations.

Fewer than two thirds (61%) of physicians knew that the HPV vaccine consists of recombinant HPV protein.

### Knowledge compared by medical specialism

The proportion of correct answers varied among the physicians according to their speciality, although the differences were not statistically significant overall (p>0.05). Overall, a higher number of correct answers came from paediatricians (65%) than from other groups.

Gynaecologists were more likely than other specialities to answer correctly that HPV is most common in women in their 30s (OD 0.12, p=0.000824), although as only 40.9% of gynaecologists answered correctly, knowledge was low over all specialties. Gyanecologists were, however, more likely to answer incorrectly that genital warts are caused by the same strains of HPV, with 65.9% giving an incorrect answer compared with 24.6% of paediatricians (OD 2.17 p=0.0246) (Table 1). This suggests that physicians’ level of knowledge is inconsistent across specialties.

For the other questions, differences between specialties were not statistically significant, though paediatricians (74.8%) and oncologists (74.2%) returned a higher rate of correct responses than family practitioners (70.9%) and gynaecologists (68.2%) for the question, “All common cervical cancers are caused by HPV” (p=0.03). There was no significant difference between the specialties in their understanding of whether people with HPV are likely to be symptomatic (they were equally incorrect) (p=0.187); that cervical cancer is one of the most common cancers amongst women, which most answered correctly (p=0.08); and whether or not HPV is a relatively common infection (p=0.643), which just over half of family physicians, oncologists and paediatricians believed to be true, and just under half of the gynaecologists (46.5%).

#### Knowledge compared by age and length of professional service

The proportion of correct answers was significantly different across the age groups (p=0.004). Respondents in the age group 45-59 years old returned a higher rate of correct answers overall. Physicians in the age group under 44 years old were more likely (69.1%) to give a correct answer for the question regarding how common HPV is (Table 1) compared with physicians in age groups 45-59 and over 60s (OD 2.56 and 5.26, p-value 0.023637 and 0.000724 respectfully) (Table 3). The 45-59 age group were also more likely to answer correctly that almost all cervical cancers are caused by HPV infection (77.8%) than those in the lower age group (68.3%, p = 0.018) and the over 60s.

The proportion of correct answers was significantly different when compared across the number of years of service. Respondents with 15–19 years of service answered questions correctly more often (50%) than those who had been in the profession for both a longer (30%) and a shorter time (44.6%) (p=0.003). This differences may be linked to mid-career physicians being more likely to still attend courses, and to engage in more educational activities than those nearing the end of their careers, providing greater opportunities for keeping up-to-date with new developments.

#### Attitudes towards HPV vaccination

The survey contained six questions that measured confidence or hesitancy to vaccines in general and the HPV vaccine in particular. The majority of the respondents (87.7%) supported the national vaccination programme for general childhood vaccination but fewer than two-thirds (63.8%) agreed that the introduction of the HPV vaccine into the national vaccination programme was appropriate. More than half (53.3%) expressed concern over the efficacy the HPV vaccine, 61.0% expressed concerns over its safety, and 58.50% agreed with the statement that the vaccine is “too new and hasn’t been around long enough”. Just under half (42.9%) agreed with the statement that 13 is too young for a child to receive the vaccine (Figure 2).

**Figure 2:**
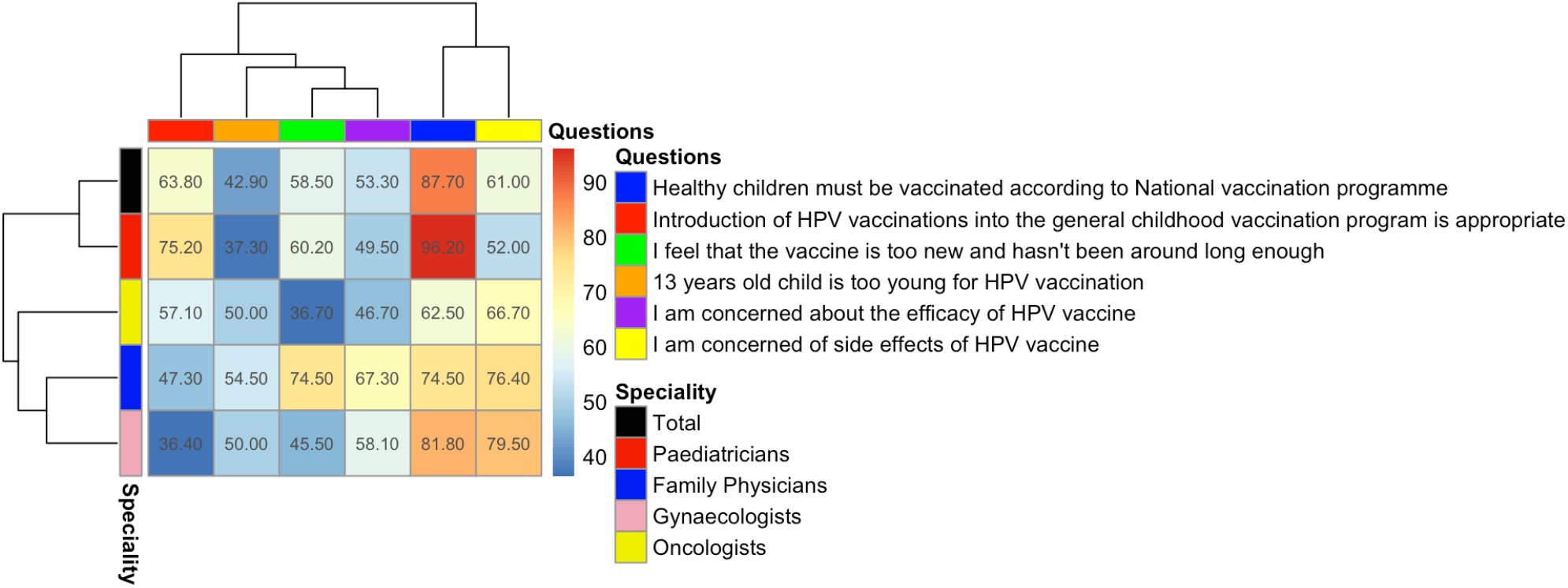
Hierarchy clustering heatmap showing opinions of physicians from different speciality to the HPV vaccine. Number in squares indicate the percentage who said yes.

#### Support for vaccination by specialty

Different specialties displayed statistically significant (p=0.000) attitudes to the introduction of the HPV vaccine into the national vaccination programme: a higher proportion of paediatricians (75.2%) agreed with it, which was statistically higher than the other groups (oncologists 57%; family physicians 47% and gynaecologists 36%). Paediatricians were more likely to agree with the need for the HPV vaccination programme (75.2%) than other specialists, which is consistent with a study from Serbia in which nearly two-thirds of the paediatricians were willing to recommend the vaccine (60.2%).^27^

The proportion of physicians concerned about side effects of the vaccine (including the risk of infertility, for which there is no medical evidence) also differed significantly among different specialties (p=0.000), with a higher percentage of family physicians and gynaecologists showing concern (respectively 76.4% and 79.5%) than paediatricians and oncologists (52% and 67%). Physicians over 60 years old showed a statistically significantly different level of concern about efficiency and the side effects than those under 60 (respectively p=0.003, p=0.002) (Tables 1 and 2).

**Table 2.**
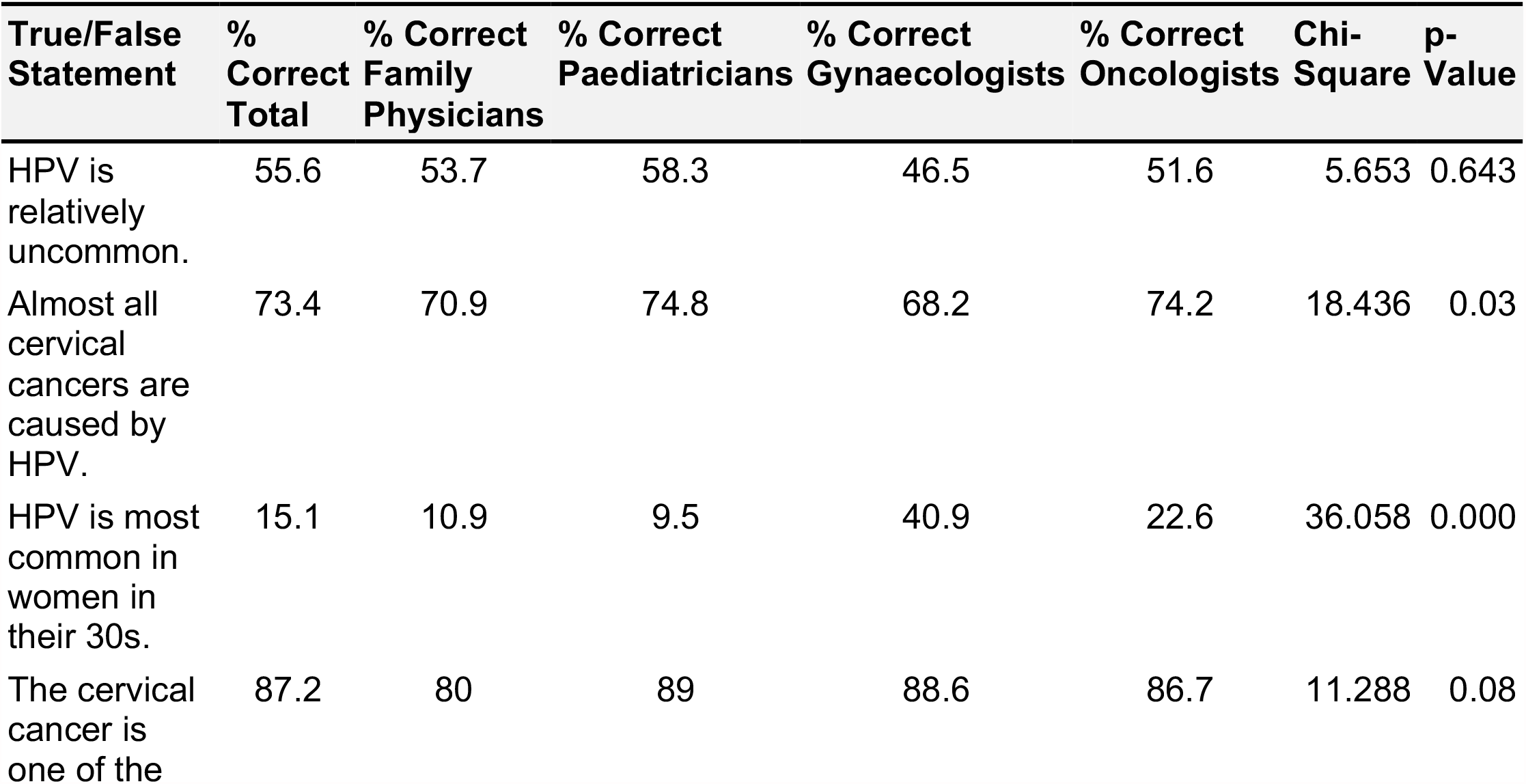
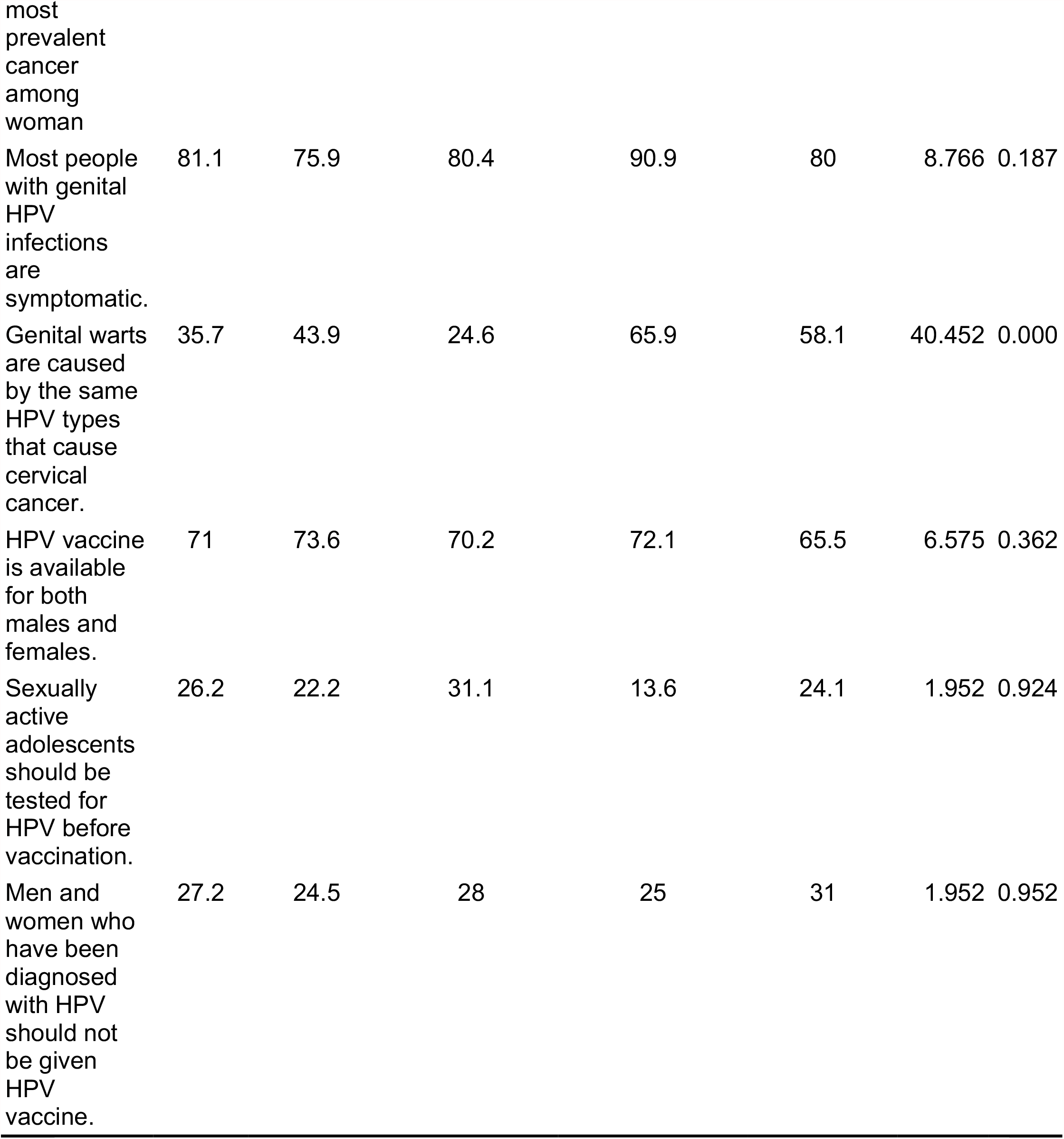
Knowledge of HPV: Comparison between physicians practicing different specialist areas.

There was no statistically significant difference in the specialty of physicians who were concerned about the newness of the vaccine (p=0.01) but there was with regard to the age at which it should be given (p=0.000 respectively). The highest percentage of physicians who were concerned that the vaccine was too new (74.5%) and that a 13-year-old was too young to receive the vaccine (54.5%) was family physicians, suggesting that this speciality is more prone to vaccine hesitancy than others, the reasons for which require further investigation.

Respondents who were concerned about efficacy were more likely to hold the opinion that the HPV vaccine is too new and “has not been around long enough” (58.8%) (Figure 2).

The proportion of physicians over the age of 60 who were concerned that 13-year-old females were too young for the vaccination was higher (55.4%) than in younger physicians (39.2%), suggesting that attitudes can differ between demographic groups. The differences in answers to this question among the different age groups was statistically significant (p=0.003).

In a final question, respondents reported that they had gained information about HPV vaccine from conferences and special events organized by the Ministry of Health, Armenia (72%), specialist literature (63%), colleagues (58%), and self-guided internet searches (57%).

## Discussion

Our study shows that there are knowledge gaps in Armenian physicians’ understanding with regards to some basic aspects of HPV infection and the HPV vaccines that is likely to influence not only the administration of HPV vaccines but also other national vaccination programmes and rollouts, including that for COVID-19, since some of the underlying concerns are likely to be generalizable. These knowledge gaps affect physicians’ understanding of disease transmission; symptoms and presentation; who will benefit from the vaccine; and vaccine safety and efficacy.

The level understanding amongst Armenian physicians with regard to the mechanisms of disease and vaccination efficacy and benefits is poor. The majority of the physicians knew that cervical cancer is one of the most prevalent types of cancer among women and that it is caused by the HPV virus but only half of the participants knew that genital warts and cervical cancer are caused by different strains of HPV and a third were unaware that the HPV vaccine is available for both females and males. This suggests that better information about vaccine-controllable diseases and their aetiology may need to be provided as part of vaccination roll-out programmes to ensure that physicians are well-informed, and their information is up to date. There may be value in more systematically embedding awareness training as part of continuing professional development programmes for healthcare workers who are likely to be in roles that can influence vaccine take-up or hesitancy.

Almost all respondents were aware of the HPV vaccine. Paediatricians were significantly more knowledgeable than other speciality groups, possibly because they were the group most likely to be dealing with the age group that is recommended for vaccination. However, the results show that knowing a vaccine is available does not always correspond to being well informed about it or the disease it can prevent. 62% of physicians were concerned about side effects of the HPV vaccine (although they were not able to specify what any such side effects might be when asked) suggesting that honest discussions around any likely side effects, including their frequency, treatment and patient outcomes may help to build healthcare practitioner confidence. In addition, 54% of the physicians were worried about the effectiveness of the HPV vaccine, which again could be addressed by in-job training and awareness that would help them to impart greater confidence to patients, who are likely to look to healthcare professionals to advise them.

The particularly low number of correct answers given by the longer-serving group (Table 1) suggests that continued professional training after qualification, to keep one’s skills up-to-date, may be somewhat lacking; although the least experienced group also scored lower than the group with mid-level experience and further study is required to understand the reasons for this.

These knowledge gaps exist despite the fact that almost all respondents reported participating in special events, such as training and seminars, and mentioned specialized literature as one of their main sources of information, suggesting that the current training and methods of information dissemination are ineffective. Further studies are warranted to understand the reasons behind these statistics, particularly as physicians can act as powerful gatekeepers whose attitudes may have ramifications for vaccine hesitancy and acceptance within the communities of patients they attend.

### Cultural assumptions and misconceptions

It is noted that cultural assumptions can deflect from the scientific basis of vaccine schedules. In Armenia, the routine early childhood immunisation coverage is estimated at 93%-97% of all children and physicians strongly support vaccination programmes amongst this age group.^28^ This study suggests that whilst early childhood vaccinations are strongly supported by most Armenian physicians (87.70%), fewer are comfortable with the inclusion of the HPV vaccine into the national vaccination programme (63.80%). The physicians had a tendency to associate HPV vaccination with the onset of sexual activity and felt that support for the vaccination of young girls was a tacit acceptance of early sexual activity. This attitude was more frequently found among physicians over 60 years old, suggesting that vaccine hesitancy may be more complex than a binary ‘yes/no’: a ‘no’ for one age cohort may become a ‘yes’ for another. However, the age of vaccine administration is not, in fact, determined by the age at which sexual activity is likely to begin: according to the WHO, the recommended priority age for HPV vaccination is 9-13 years old because this is the age at which the immune system response is highest and produces the necessary level of antibodies and immune-system memory to protect against HPV infection for life, not because this is the age at which children may become sexually active. It is important for preteens to get all three doses long before any sexual activity begins.^29^ This should not affect the administration of vaccines for COVID-19 but does show how cultural attitudes and beliefs intersect with scientific evidence to build acceptance or resistance to vaccination. Vaccine confidence or hesitancy can be based on sociocultural factors as well as medical science.

Controversies among health professionals relating to the inclusion of HPV in national immunization schedules has also been described in Spain, where 89% of health professionals were aware of the relationship between HPV infection and cervical cancer but 65.7% resist its introduction into vaccine schedules, arguing that there is no data on its long-term effectiveness.^25^

The Armenian physicians also expressed concern over the newness of the Gardasil vaccine (from 36.7% amongst oncologists to 74.5% in family physicians; 58.5% across all specialities). This is particularly concerning with regard to COVID-19, as all the vaccines for this disease (and likely for future emerging diseases) are new and have been developed more quickly than is usual for new pharmaceuticals. This may require targeted education to help healthcare providers understand the development and approvals process for these new vaccines to give assurance that it is the paperwork, funding mechanisms, and administration around the clinical trials and approvals that has been speeded up, not the safety aspects of trials^24^. This in turn this could be used to reassure patients that new vaccines are ‘safe’.

By paying more attention to the disconnect between the scientific basis for the decision to vaccinate and the cultural assumptions against vaccination at this particular age, we may be able start to explore how such attitudes can be changed. It also illustrates that by not being aware of such assumptions and failing to take time to address them in awareness information and training given to professionals responsible for administering the vaccine, vaccine confidence can be undermined by misconceptions and cultural perceptions that may prove harder to dispel later on.

### Vaccine hesitancy may not be universal

The different levels of confidence in the national childhood vaccination programme compared with the introduction of the HPV vaccination highlight that different vaccines can be considered independently of one another. Vaccine hesitancy may not be universal to all vaccines nor have the same underlying concerns with regard to each vaccine. Understanding this more fully would benefit from further research to help determine how different forms of hesitancy can be effectively challenged and what steps might be taken in advance to stop such hesitancy from becoming widespread within certain communities.

Further studies could also usefully determine the extent to which physicians’ poor understanding, misconceptions and hesitancy may influence their decision to administer vaccines and their ability to sufficiently inform patients of vaccine benefits.

## Conclusion

This study shows that the concerns and worries that influence negative attitudes towards vaccines, and which may prevent physicians from recommending vaccination, are complex. Reasons may differ for different diseases and may be influenced by different underlying factors, all of which suggest the need for stronger vaccine-confidence awareness training that helps physicians – and by extension, to the patients they advise – to navigate these concerns by providing up-to-date and comprehensive information that will enhance confidence and dispel unwarranted concerns. The role of the media, and its portrayal of HPV vaccination is likely to have some influence in this process and should be considered within future research programmes.

Differences in the approach of different physicians in Armenia to HPV vaccination was evident in this study, with different approaches noted based on their age, length of service, specialty and cultural assumptions. Overall, however, the study merely highlights gaps our complete understanding of vaccine hesitancy and how this may vary between different vaccines and for the prevention of different diseases; it does not address them. In this regard, priority consideration should be given to further study and, in the short term, to developing a programme of continuous education on all aspects of vaccination development, safety, efficacy and societal value, both for HPV infections and for other newly developed or introduced vaccines. Further research is needed to fully investigate all underlying reasons for vaccine hesitancy, and to develop interventions to address this and improve confidence.

## Data Availability

The data used for analyses is presented in the paper.

## Authors’ contributions

MH, GG and MMT data collection, methodology, conceptualisation; ARB, RC data curation and data analysis; JC writing, review and editing; AWK review and editing; HVM data curation, writing, editing and review. All authors contributed to the article and approved the submitted version.

## Acknowledgements

The authors thank the National Center for Disease Control and Prevention of Ministry of Health of Republic of Armenia and Yerevan State Medical University for support the implementation of the research.

## Ethics

This study was approved by the Ethics Committee of Yerevan State Medical University after Mkhitar Heratsi (YSMU 14.06.2016/No 10).

## Availability of data and material

The data used for analyses is presented in the paper.

