## Supplementary material for "The Knowledge and Attitude of Physicians Regarding Vaccinations in Yerevan, Armenia: Challenges for COVID-19": Table S1

|  | HPV is relatively uncommon. | | | Almost all cervical cancers are caused by HPV | | | HPV is most common in women in their 30s | | | Cervical cancer is one of the most prevalent cancer among woman | | | Most people with genital HPV are symptomatic | | | Genital warts are caused by the same HPV types that cause cervical cancer. | | | Sexually active adolescent should be tested before HPV vaccination | | | HPV vaccine is available for both males and females. | | | Men and women who have been diagnosed with HPV should not be given HPV vaccine. | | |
| --- | --- | --- | --- | --- | --- | --- | --- | --- | --- | --- | --- | --- | --- | --- | --- | --- | --- | --- | --- | --- | --- | --- | --- | --- | --- | --- | --- |
|  | OR | 95% CI | p value | OR | 95% CI | p value | OR | 95% CI | p value | OR | 95% CI | p value | OR | 95% CI | p value | OR | 95% CI | p value | OR | 95% CI | p value | OR | 95% CI | p value | OR | 95% CI | p value |
| Speciality |  |  |  |  |  |  |  |  |  |  |  |  |  |  |  |  |  |  |  |  |  |  |  |  |  |  |  |
| Family Physicians | 1.00 | - | - | 1.00 | - | - | 1.00 | - | - | 1.00 | - | - | 1.00 | - | - | 1.00 | - | - | 1.00 | - | - | 1.00 | - | - | 1.00 | - | - |
| Gynaecology | 1.60 | 0.67 -3.88 | 0.287346 | 1.07 | 0.41 -2.78 | 0.8843 | 0.12 | 0.03 -0.39 | 0.000824 | 0.37 | 0.09 - 1.24 | 0.1258 | 0.37 | 0.09 -1.24 | 0.18321 | 0.42 | 0.17- 1.00 | 0.0538 | 1.96 | 0.65 -6.40 | 0.241 | 1.18 | 0.46- 3.04 | 0.7307 | 1.01 | 0.38 - 2.69 | 0.987 |
| Oncology | 1.15 | 0.42- 3.16 | 0.790389 | 0.83 | 0.25 -2.52 | 0.7443 | 0.28 | 0.06 - 1.11 | 0.075541 | 0.53 | 0.11- 2.00 | 0.3763 | 0.53 | 0.11- 2.00 | 0.96611 | 0.54 | 0.20- 1.43 | 0.2179 | 0.80 | 0.25 -2.68 | 0.7135 | 1.50 | 0.51 - 4.32 | 0.4492 | 0.65 | 0.22 - 1.95 | 0.437 |
| Paediatrician | 0.93 | 0.48 -1.82 | 0.834092 | 0.79 | 0.39 -1.68 | 0.5309 | 0.86 | 0.23 - 2.58 | 0.808196 | 0.50 | 0.22 1.19 | 0.1061 | 0.50 | 0.22- 1.19 | 0.52803 | 2.17 | 1.10- 4.26 | 0.0246 | 0.81 | 0.36- 1.72 | 0.5974 | 1.19 | 0.59- 2.53 | 0.6415 | 0.81 | 0.37 - 1.66 | 0.57 |
| Age |  |  |  |  |  |  |  |  |  |  |  |  |  |  |  |  |  |  |  |  |  |  |  |  |  |  |  |
| less than 29 | 1.82 | 0.41 -7.53 | 0.3656 | 2.55 | 0.62-10.55 | 0.1871 | 6.32 | 0.64 -164.50 | 0.168805 | 0.28 | 0.01- 2.26 | 0.3012 | 0.28 | 0.01- 2.26 | 0.14421 | 1.27 | 0.30 -6.48 | 0.7599 | 0.73 | 0.15- 4.17 | 0.708 | 0.86 | 0.18 - 3.55 | 0.8378 | 0.72 | 0.1631 - 3.37 | 0.661 |
| 30-44 | 1.00 | - | - | 1.00 | - | - | 1.00 | - | - | 1.00 | - | - | 1.00 | - | - | 1.00 | - | - | 1.00 | - | - | 1.00 | - | - | - | - | - |
| 45-59 | 2.59 | 1.06-5.31 | 0.023637 | 0.70 | 0.27 -1.83 | 0.4662 | 0.96 | 0.32 -2.79 | 0.938792 | 0.88 | 0.27 -3.15 | 0.836 | 0.88 | 0.27 -3.15 | 0.7172 | 0.98 | 0.42- 2.22 | 0.9545 | 0.52 | 0.21- 1.24 | 0.1498 | 0.80 | 0.34-1.92 | 0.6123 | 0.99 | 0.43- 2.22 | 0.974 |
| Over 60 | 5.26 | 2.04-14.07 | 0.000724 | 0.82 | 0.28 -2.41 | 0.7127 | 1.35 | 0.34 -5.65 | 0.671597 | 0.66 | 0.16 - 2.83 | 0.5719 | 0.66 | 0.16- 2.83 | 0.60695 | 0.99 | 0.37- 2.67 | 0.9907 | 2.07 | 0.66- 6.70 | 0.2135 | 1.06 | 0.39 - 2.88 | 0.9131 | 1.35 | 0.50 - 3.68 | 0.558 |
| Experience |  |  |  |  |  |  |  |  |  |  |  |  |  |  |  |  |  |  |  |  |  |  |  |  |  |  |  |
| Less than 5 | 0.91 | 0.18 -4.18 | 0.902381 | 2.56 | 0.52-13.18 | 0.2483 | 0.42 | 0.06 -2.91 | 0.378948 | 5.45 | 0.65 - 56.15 | 0.1224 | 5.45 | 0.65 - 56.15 | 0.72554 | 1.20 | 0.25- 5.74 | 0.8201 | 2.56 | 0.33 - 5.02 | 0.3331 | 1.88 | 0.38 - 9.19 | 0.4306 | 2.05 | 0.41 - 12.87 | 0.403 |
| 5 to 9 | 0.57 | 0.14 -2.08 | 0.403586 | 1.80 | 0.44-7.71 | 0.4148 | 2.04 | 0.40 -12.58 | 0.407467 | 1.49 | 0.16 -14.02 | 0.7123 | 1.49 | 0.16- 14.02 | 0.30059 | 0.83 | 0.24- 2.92 | 0.7695 | 1.24 | 0.33 - 5.02 | 0.7491 | 1.74 | 0.47 - 6.53 | 0.4027 | 1.45 | 0.40 - 5.70 | 0.583 |
| 10 to 14 | 1.00 | - | - | 1.00 | - | - | 1.00 | - | - | 1.00 | - | - | 1.00 | - | - | 1.00 | - | - | 1.00 | - | - | 1.00 | - | - | 1.00 | - | - |
| 15 to 19 | 1.03 | 0.37 -2.89 | 0.955437 | 0.98 | 0.27 -3.84 | 0.978 | 1.20 | 0.33 - 4.25 | 0.783078 | 1.89 | 0.35 - 14.72 | 0.4852 | 1.89 | 0.35- 14.72 | 0.27424 | 0.80 | 0.28- 2.26 | 0.6708 | 1.55 | 0.51 - 4.66 | 0.4362 | 1.11 | 0.35 - 3.64 | 0.8584 | 0.78 | 0.27 - 2.18 | 0.644 |
| More than 20 | 0.71 | 0.28 -1.82 | 0.475597 | 1.94 | 0.64 -7.01 | 0.2693 | 1.96 | 0.53 - 6.54 | 0.290064 | 2.49 | 0.55 -18.75 | 0.2932 | 2.49 | 0.55- 18.75 | 0.13082 | 1.72 | 0.64 - 4.51 | 0.2724 | 1.61 | 0.59 - 4.28 | 0.3427 | 1.41 | 0.52 - 4.30 | 0.5212 | 1.27 | 0.47 - 3.27 | 0.624 |

**Table S1. Multivariate logistic regression analysis of respondents’ awareness of HPV and it’s vaccine in Armenia**

OD = odd’s ratio, CI= confident interval ,Highlighted p value >.0.05

t
